# Evidence for Cytokine Dysregulation in Schizophrenia Spectrum Disorders: A Comparison of Cerebrospinal Fluid and Blood Samples

**DOI:** 10.1101/2023.01.25.23285017

**Authors:** Juan A. Gallego, Robert K. McNamara, Santiago Castaneda, Laura D. Jimenez, Santiago Alvarez-Lesmes, Emily A. Blanco, Todd Lencz, Anil K. Malhotra

## Abstract

**Introduction:** Abnormalities in immune function have been described in schizophrenia. Cytokines, key signaling molecules of the immune system, have been investigated in peripheral blood samples of patients with schizophrenia though few studies have investigated cytokines in cerebrospinal fluid (CSF). Moreover, very few studies with schizophrenia subjects have investigated the relationships between CSF and blood cytokine levels or investigated sex differences.

**Methods:** In this cross-sectional study, subjects with schizophrenia spectrum disorder (SSD) diagnosis and healthy volunteers (HV) underwent a lumbar puncture, a blood draw, and psychopathology ratings. CSF and plasma levels of IL-1ß, IL-2, IL-4, IL-6, IL-8 and TNFα were determined by enzyme-linked immunosorbent assay (ELISA) and a high-sensitivity MilliplexTM Multiplex kit. Cytokine levels were square root transformed and compared between both groups using bivariate tests and a multivariate regression analysis. CSF and plasma cytokine levels were compared as were the effects of sex.

**Results:** Thirty SSD and 23 HV were included in the study. In bivariate analysis, CSF TNFα and IL-4 levels were significantly increased in SSD subjects compared to HV. Multivariate regression analysis also showed increases in TNFα and IL-4 after adjusting for sex, age and body mass index. There were no significant differences in plasma cytokine levels between groups, and there were no correlations between CSF and plasma cytokine levels. CSF levels of TNFα and IL-4 were negatively correlated with speed of processing. Stratified analysis by sex showed significantly increased levels of TNFα and IL-4 in SSD vs. HV in males but not in females, although sample sizes were small after stratification.

**Discussion:** The present findings provide evidence of dysregulation of TNFα and IL-4 in CSF of schizophrenia and suggest that peripheral cytokine levels are not reflective of CSF levels. We also provide further evidence of the association between cytokine levels and impairment of certain cognitive domains. These results identify elevated CSF TNFα levels as a potential biomarker relevant to the neuropathophysiology of schizophrenia.

## 1. Introduction

One of the leading hypotheses in the pathophysiology of schizophrenia is the dysregulation of the immune system. Several molecules of the immune system have been studied as part of this hypothesis; however, cytokines have been prominently investigated given their critical role in the immune response (Salvador et al., 2021). Cytokines are a group of small proteins secreted by immune and nonimmune cells and are key members in the regulation of the innate and adaptive immune system (Parkin et al., 2001). Cytokines, which bind to both receptors in target cells and soluble receptors, cross the blood brain barrier via saturable transport systems (Banks et al., 1995). Although other molecules involved in the immune system function have been studied in patients with schizophrenia, including complement component 4 (C4) (Sekar et al., 2016, Gallego et al., 2021), C-reactive protein (Fond et al., 2018), and oxidative stress molecules (Flatow et al., 2013; Upthegrove et al., 2020), additional research is needed to better characterize their role of cytokines in central pathophysiological processes.

Increased levels of pro-inflammatory cytokines have been described in peripheral blood and cerebrospinal fluid (CSF) samples of patients with schizophrenia, as reported by individual studies, such as those by C. G. McAllister et al. 1995, Garver et al. 2003, Pandey et al. 2015, Boerrigter et al. 2017, and as summarized in meta-analyses by our group (Gallego et al., 2018) and other groups (Miller et al., 2011; Wang et al., 2018). Despite this relatively large body of work, several gaps still exist in the literature. First, the majority of studies examined cytokine levels in peripheral blood samples (Wang et al., 2018) whereas few have been conducted using CSF samples (Gallego et al., 2018), even though schizophrenia is considered a brain disorder and cytokines require saturable transports to cross the blood brain barrier (Banks et al., 1995).

Second, several cytokines have been either understudied or underreported in CSF studies. For example, TNFα, despite being a major regulator of inflammatory responses (Kalliolas et al., 2016), has only been reported in CSF in a small number of studies and results have been inconsistent (Barak et al., 1996; Maxeiner et al., 2014; Zhu et al., 2016). Importantly, the sample sizes of those studies were small, and the only study that used a healthy population as a control group did not find any measurable levels in either schizophrenia patients or healthy volunteers (Barak et al., 1996). Third, despite the critical importance of determining whether peripheral blood cytokine levels accurately reflect central cytokine levels, very few studies have directly investigated the relationship between blood and CSF cytokine levels in schizophrenia patients. Two studies did not find a correlation between blood and CSF IL-6 (Sasayama et al., 2013) or IL-1B (Katila et al., 1994) levels, whereas another study found a correlation between CSF and blood in IL-6 levels in a small sample 11 of recent onset schizophrenia patients and 12 healthy controls (Coughlin et al., 2016). Other studies measured cytokines in both CSF and blood samples of patients with schizophrenia, but correlations were not computed (McAllister et al., 1995, Maxeiner et al., 2014). Therefore, additional studies are needed to better characterize peripheral and central cytokine levels in patients with schizophrenia.

Differences in immune response have been demonstrated between males and females. Females reportedly have a higher innate and adaptive immune response compared to males as demonstrated by the higher susceptibility of females to suffer from autoimmune and inflammatory diseases, higher antibody response to certain vaccines, higher basal immunoglobulin levels and higher B cell numbers (Klein et al., 2016), with the immunosuppressive effect of androgens being one of the factors implicated in these differences. Despite this, few studies have reported the impact of sex on blood and CSF cytokine levels in patients with schizophrenia (Gaver et al., 2003, O’Connell et al., 2014, Schwieler et al., 2015; Lee et al., 2019). Two studies found elevated peripheral blood levels of IL-1ß, IL-8, IL-17, IL-23 and TNFα (O’Connell et al., 2014) and IL-6 (Lee et al., 2019) in female patients compared to male patients. Garver and colleagues (2003) found no sex differences in IL-6 in CSF whereas Schwieler and colleagues (2015) found higher CSF IL-6 levels in both males and female patients compared with same-sex controls and overall higher levels in females compared to males. Therefore, additional studies are needed to better characterize sex effects on peripheral and central cytokine levels in patients with schizophrenia.

To address these gaps, the present study investigated a panel of cytokines in both CSF and plasma from 33 patients with schizophrenia spectrum disorders (SSD) and 23 healthy volunteers (HV). Our primary goals were: 1) to determine cytokine levels in both plasma and CSF in SSD patients compared with HV; 2) to investigate associations between CSF and plasma cytokines levels, and 3) to investigate the impact of sex on CSF cytokine levels.

## 2. Material and Methods

### 2.1. Inclusion and Exclusion Criteria

Patients were included in the study if they (1) fulfilled DSM-IV criteria for schizophrenia, schizophreniform disorder, schizoaffective disorder, or Psychosis Not Otherwise Specified (NOS); (2) were 15-59 years old, and (3) were willing and capable of providing informed consent. Subjects were excluded from participation if they (1) were prescribed an anti-coagulant; (2) had a history of an organic brain disorder, (3) had a clinically significant thrombocytopenia or coagulopathy based on screening blood tests that prevented them for having a lumbar puncture, or (4) had a diagnosis of a substance-induced psychotic disorder. Healthy controls were also recruited and fulfilled the same inclusion criteria except for the diagnosis’s criterion. They were excluded if they had an Axis I diagnosis or if they had a first-degree relative with a known or suspected Axis I disorder, based on family history questionnaire.

### 2.2. Recruitment and Consent

SSD patients were recruited from the inpatient and outpatient departments at The Zucker Hillside Hospital, a tertiary psychiatric hospital from Northwell Health in Glen Oaks, NY. HV were recruited from the general population via word of mouth, newspaper and internet advertisements, and poster flyers. All subjects provided written informed consent to a protocol approved by the Institutional Review Board of the Northwell Health System. After providing informed consent subjects provided urine for a urine toxicology test, underwent baseline ratings, including demographic information. Psychopathology ratings were administered using the Brief Psychiatric Rating Scale (BPRS), the schedule for assessment of negative symptoms (SANS) and the Clinical Global Impression severity index (CGI-S). A neuropsychological assessment was administered using the MATRICS Consensus Cognitive Battery (MCCB).

### 2.3. CSF and blood collection

Subjects underwent a lumbar puncture, performed using a standard technique with a 25 gauge, Whitacre point spinal needle after subcutaneous lidocaine was applied. The procedure was conducted with patient sitting up and all procedures took place at 2 pm. Approximately 15–25 cc of CSF were obtained from each subject. The first 2-3 mls of CSF were sent for routine testing (cell count, proteins, glucose, and VDRL). CSF samples with macroscopic blood as consequence of traumatic procedures were not included in these analyses. A blood draw to obtain 10 cc of peripheral blood was obtained prior to the lumbar puncture procedure. Both CSF and blood samples were centrifuged to remove cells in the case of CSF samples or to isolate plasma in the case of blood. Both sample types were flash frozen with liquid nitrogen and stored at −80 Celsius at the biorepository of the Feinstein Institute for Medical Research, the research institute for Northwell Health located in Manhasset, NY. CSF and plasma samples were sent for cytokine analyses at the University of Cincinnati.

### 2.4. CSF and plasma cytokine levels

CSF and plasma cytokine concentrations were determined in duplicate by enzyme-linked immunosorbent assay (ELISA) using MilliplexTM Multiplex kits (Millipore, Billerica, MA) according to manufacturer’s protocol. Briefly, using a 96 well black plate 25 μL sample was incubated with 25 μL antibody coated beads at 4°C on a plate shaker overnight. Plates were then washed 2 times on a vacuum apparatus and 25 μL of secondary antibody was added and incubated at room temperature for 1 hour while shaking. Strept-avidin-RPE (25 μL) was then added and incubated with the secondary antibody for 30 minutes at room temperature with shaking. Plates were then washed 2 times and 100 μL of sheath fluid added. Plates were shaken for 5 minutes and then read using Luminex technology on the Bio-PlexTM (Bio-Rad, Hercules, CA). Concentrations were calculated from standard curves using recombinant proteins and expressed in pg/ml. Samples were processed by a technician blinded to group assignment. The lower limits of detection were 1.857pg/ml for TNFα, 0.496 pg/ml for IL-1ß, 7.866 pg/ml for IL-2, 8.635 pg/ml for IL-4, 2.773 pg/ml for IL-6 and 0.318 pg/ml for IL-8. For samples with levels below the detection limit we imputed the data with a value that corresponded to half the detection limit to reflect the median value between 0 and the detection limit. Accordingly, 20/53 data points were imputed for IL-4 and 4/53 were imputed for TNFα. No data points were imputed for IL-6 and IL-8. Lastly, IL-1ß and IL-2 were not included in the analysis since most of the data points fell below the limit of detection.

### 2.5. Statistical Analysis

Baseline characteristics between patients and healthy volunteers were compared using t-tests for normally distributed continuous variables or Wilcoxon rank sum tests for continuous variables not normally distributed. Raw cytokine values did not follow a normal distribution and values were square-root transformed. This transformation was selected above others given that it provided the most appropriate normal distribution as tested by the Ladder and Gladder commands on Stata 15. Subsequent analyses were conducted using square root transformed data. To aid in interpretation, we will report untransformed data to help with interpretation of group differences, however, all p-values were calculated using square-root transformed values. Cytokine comparisons between SSD patients and HV were conducted using t-tests and pairwise Pearson correlation coefficients were calculated to determine associations between cytokine levels in CSF and plasma with psychopathology measures and neurocognition. To determine the association with case status as the dependent variable (patient vs. healthy control), cytokines were entered into a logistic regression model along with sex, age, and BMI given their know effects as potential confounding factors of cytokine levels. Correlation analyses examining associations between significant cytokines and other continuous variables such as age, BMI and MCCB domains were conducted. In addition, we computed correlations between CSF and plasma levels for all individual cytokines. These correlation analyses were exploratory and not adjusted for multiple comparisons due to limitations secondary to the limited sample size of our study. To determine the impact of sex in cytokine levels, we conducted cytokine analysis stratified by sex.

## 3. Results

### 3.1. Subjects

Thirty SSD patients and 23 HV were included in the analysis. There were no significant differences between SSD patients and HV in sex, age or race, and SSD patients had higher body mass index values (mean: 30.0, SD=8.1 vs. mean: 26.3, SD=2.9, p=0.04) and lower MCCB composite scores (mean=25.1, SD=16.1, p<0.0001) compared to healthy volunteers (Table 1)

**Table 1.**
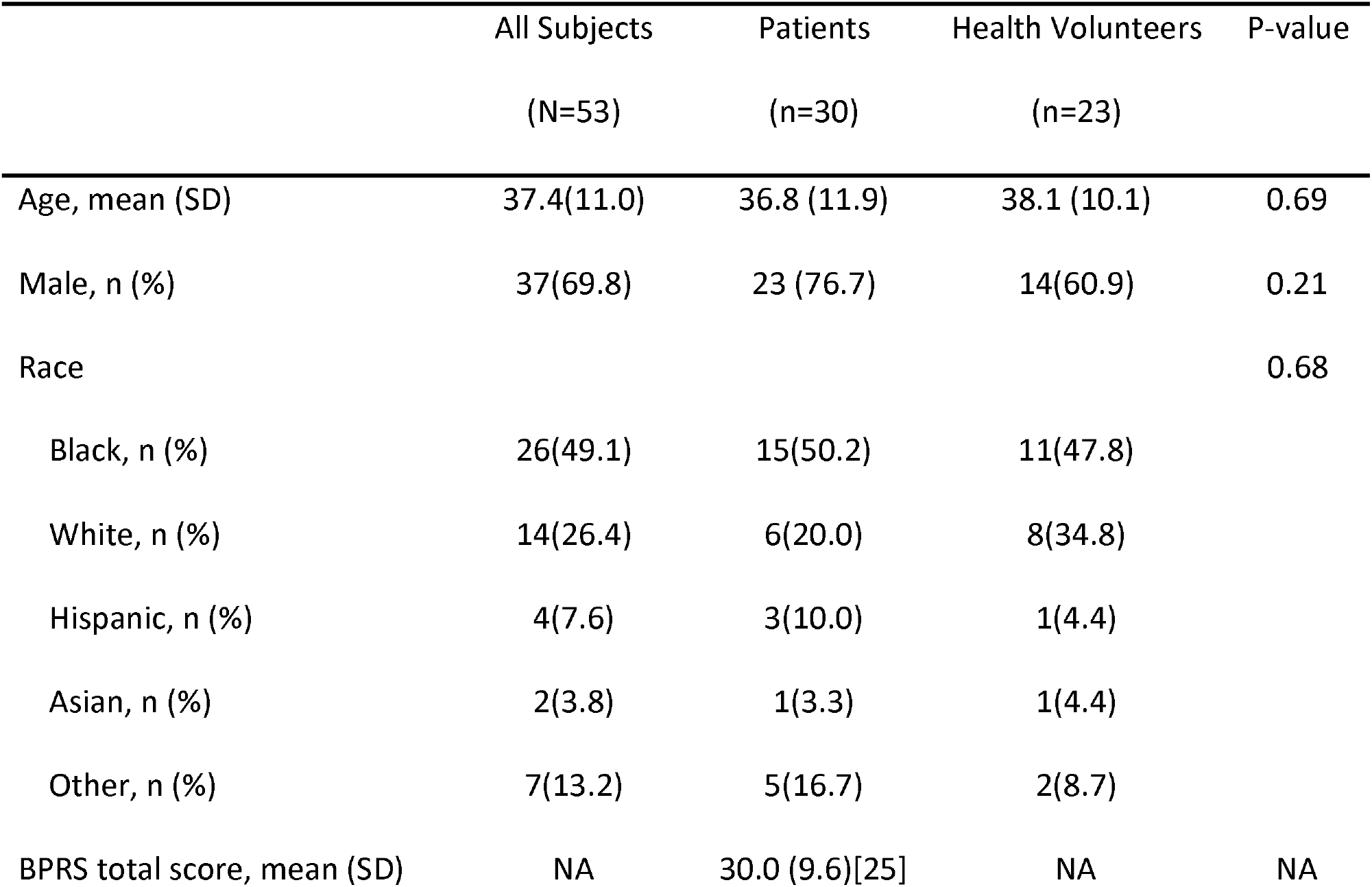
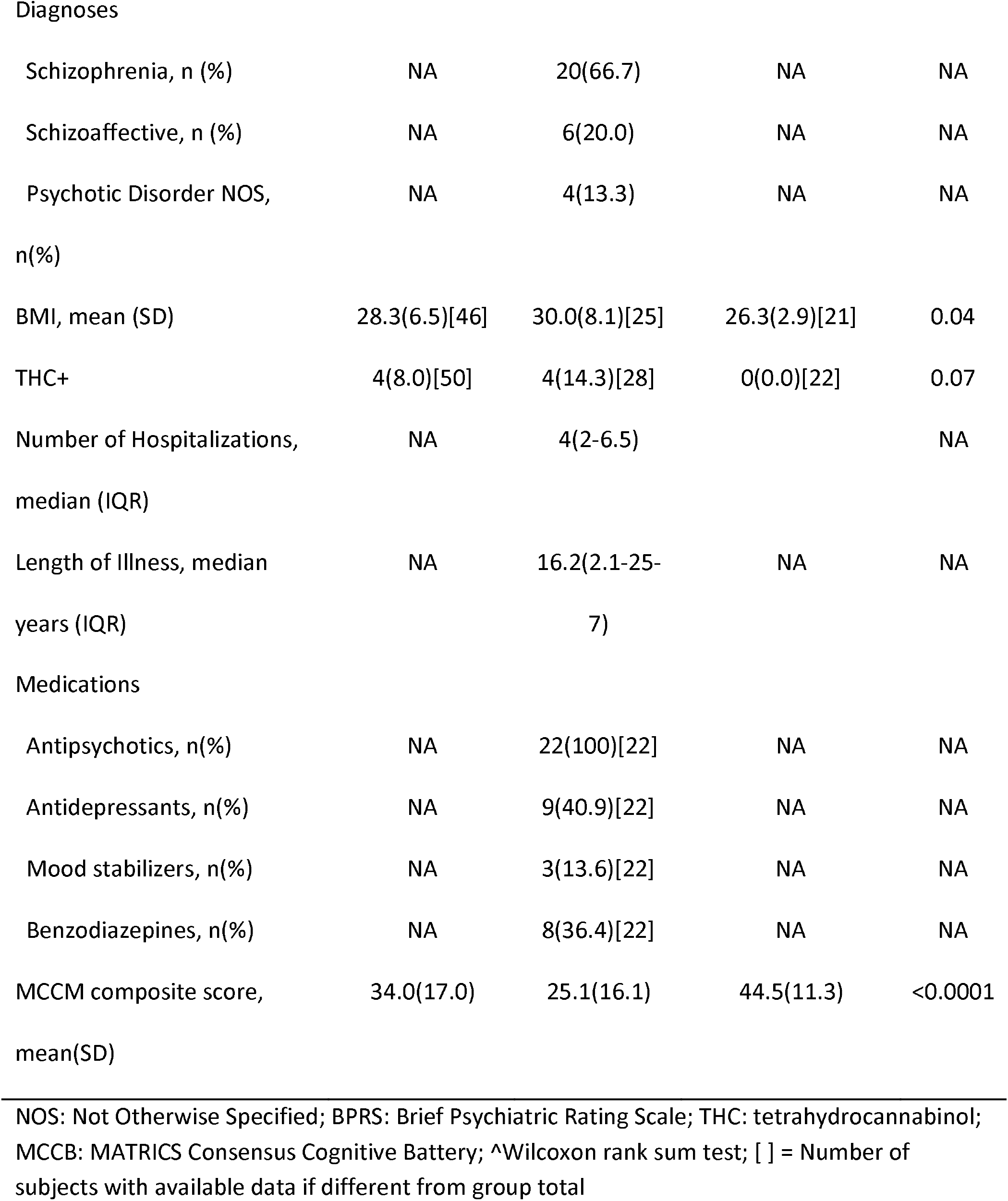
Demographic and clinical characteristics

|  | All Subjects<br>(N=53) | Patients<br>(n=30) | Health Volunteers<br>(n=23) | P-value |
| --- | --- | --- | --- | --- |
| Age, mean (SD) | 37.4(11.0) | 36.8 (11.9) | 38.1 (10.1) | 0.69 |
| Male, n (%) | 37(69.8) | 23 (76.7) | 14(60.9) | 0.21 |
| Race |  |  |  | 0.68 |
| Black, n (%) | 26(49.1) | 15(50.2) | 11(47.8) |  |
| White, n (%) | 14(26.4) | 6(20.0) | 8(34.8) |  |
| Hispanic, n (%) | 4(7.6) | 3(10.0) | 1(4.4) |  |
| Asian, n (%) | 2(3.8) | 1(3.3) | 1(4.4) |  |
| Other, n (%) | 7(13.2) | 5(16.7) | 2(8.7) |  |
| BPRS total score, mean (SD) | NA | 30.0 (9.6)[25] | NA | NA |
| Diagnoses |  |  |  |  |
| Schizophrenia, n (%) | NA | 20(66.7) | NA | NA |
| Schizoaffective, n (%) | NA | 6(20.0) | NA | NA |
| Psychotic Disorder NOS,<br>n(%) | NA | 4(13.3) | NA | NA |
| BMI, mean (SD) | 28.3(6.5)[46] | 30.0(8.1)[25] | 26.3(2.9)[21] | 0.04 |
| THC+ | 4(8.0)[50] | 4(14.3)[28] | 0(0.0)[22] | 0.07 |
| Number of Hospitalizations,<br>median (IQR) | NA | 4(2-6.5) |  | NA |
| Length of Illness, median<br>years (IQR) | NA | 16.2(2.1-25-7) | NA | NA |
| Medications |  |  |  |  |
| Antipsychotics, n(%) | NA | 22(100)[22] | NA | NA |
| Antidepressants, n(%) | NA | 9(40.9)[22] | NA | NA |
| Mood stabilizers, n(%) | NA | 3(13.6)[22] | NA | NA |
| Benzodiazepines, n(%) | NA | 8(36.4)[22] | NA | NA |
| MCCM composite score,<br>mean(SD) | 34.0(17.0) | 25.1(16.1) | 44.5(11.3) | <0.0001 |

NOS: Not Otherwise Specified; BPRS: Brief Psychiatric Rating Scale; THC: tetrahydrocannabinol; MCCB: MATRICS Consensus Cognitive Battery; ^Wilcoxon rank sum test; [ ] = Number of subjects with available data if different from group total

### 3.2. Cerebrospinal Fluid Analysis

#### 3.2.1. Cytokine levels in CSF by group

Bivariate analysis showed that TNFα levels were significantly higher in SSD patients compared to HV (mean=6.91 pg/mL, SD=0.52 vs 3.76 pg/mL SD 0.53; p=0.0001, Cohen’s d=1.16). Similarly, IL-4 levels were higher in patients compared to HV (median=8.54, IQR: 6.79-15.69 vs median=4.32, IQR: 4.32-8.54, p= 0.004) (Figure 1). We did not find statistically significant differences between groups in IL-6, or IL-8 levels (Suppl. Fig 1).

**Figure 1.**
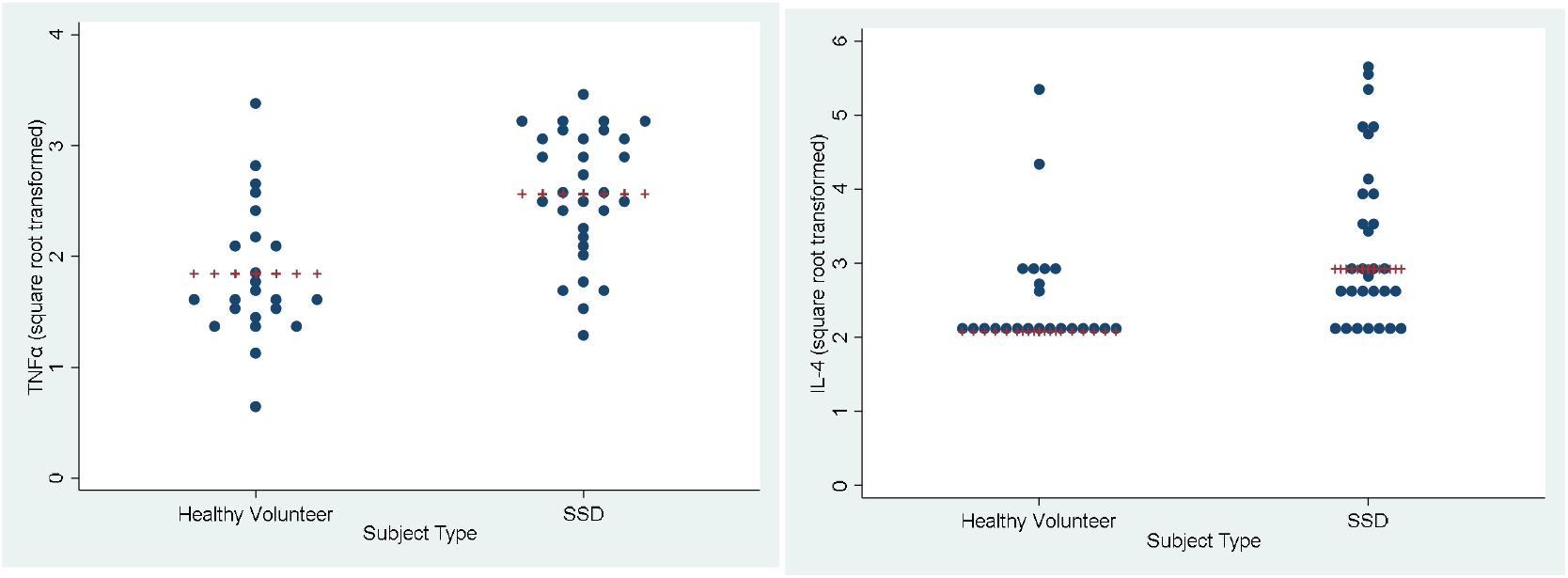
CSF levels of TNFα and IL-4 in SSD vs. HV

#### 3.2.2. Correlations between CSF cytokine levels and other measures in all subjects

In all subjects, there were no correlations between CSF TNFα and age (n=53, r=0.004, p=0.98) or BMI (n=46, r=0.09, p=0.53). A negative correlation was detected between CSF TNFα and speed of processing (n=49, r=-0.4, p=0.0024). CSF IL-4 levels were correlated with the MCCB composite score (n=48, rho=-0.32, p=0.025), the speed of processing domain of the MCCB (n=49, Spearman’s rho=-0.41, p=0.0037), the alogia domain of the SANS (n=19, rho=0.53, p=0.02), and the conceptual disorganization item of the BPRS (n=25, rho=0.45, p=0.026).

#### 3.2.3. Correlations between CSF cytokines and other measures by subject group

In SSD patients, there were no correlations between CSF TNFα and any other variables, including age, BMI, BPRS total score, SANS domains, MCCB domains, number of hospitalizations, length of illness, chlorpromazine equivalents, and CGI severity (data not shown). In HV, the attention vigilance domain was nominally correlated with CSF TNFα (n=22, r=0.44, p=0.04). There were no correlations with age, BMI, antipsychotic dose (chlorpromazine equivalents) or any other MCCB domains.

#### 3.2.4. Sex effects

CSF TNFα levels were higher in SSD compared to HV in both males (n=37) (mean: 7.06, SD: 0.63 vs. mean: 3.43, SD=0.54) and females (n=16) (mean: 6.44, SD: 0.88 vs. mean: 4.26, SD: 1.09). However, this increase was only statistically significant in males (p=0.0003) but not in females (p=0.16) (Figure 2). A similar effect was seen for CSF IL-4 levels with a statistically significant difference between SSD vs. HV in males (n=23, median=2.92, IQR: 2.08-4.11 vs. n=14, median=2.08, IQR=2.08-2.92, p=0.031) but not in females (n=7, median=2.61, IQR: 2-61-3.96 vs. n=9, median=2.08, IQR: 2:08-2:08, p=0.054).

**Figure 2.**
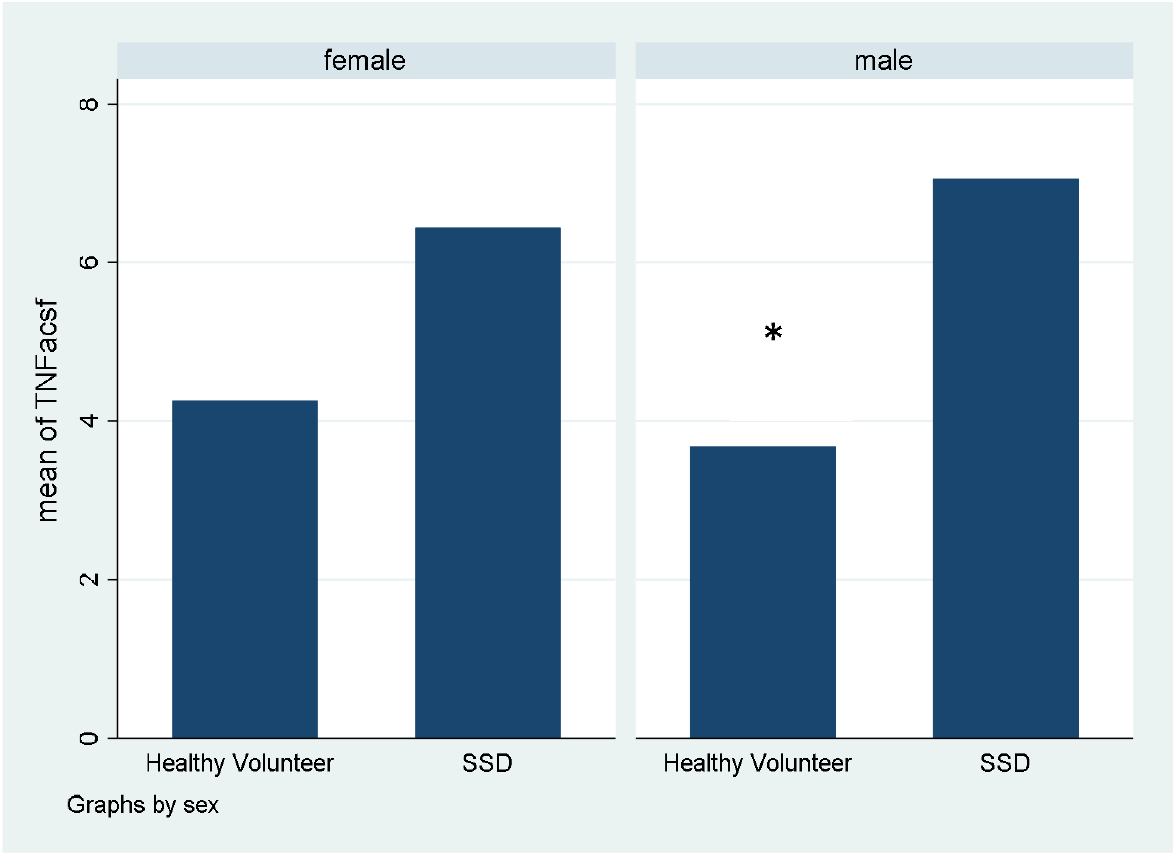
CSF TNFα levels by sex between SSD vs. HV *p=0.0003

#### 3.2.5. Multivariable regression analysis

In a series of multivariable linear regression analysis using individual cytokines as the dependent variable, an SSD diagnosis was associated with TNFα (p=0.001), and IL-4 (p=0.039) after adjusting for sex, age and BMI (Table 2). Relatedly, in a manual backwards elimination multivariable logistic regression analysis using SSD as the dependent variable and IL-4 and TNFα as independent variables along with sex, age and BMI showed that only TNFα was significantly associated with SSD (N=46, OR:5.59, 95%CI: 1.72-18.21, p=0.004) whereas IL-4 dropped from the model. In this analysis, the interaction of TNFα and sex was not statistically significant (p=0.66). Stratified analysis by sex showed the same findings described in the bivariate analysis. TNFα was significantly associated with SSD in males (N=32, OR:7.03, 95%CI: 1.46-33.89, p=0.015) but not in females (N=14, OR=4.82, 95%CI: 0=4.82, 95%CI: 056-41.44, p=0.15) when adjusting for age and BMI.

**Table 2.** CSF Cytokine levels - Multivariate regression analysis

| **TNFα** |  |  |  |
| --- | --- | --- | --- |
|  | *$\beta$ Coefficient* | *Standard Error* | *p-value* |
| SSD diagnosis | 0.70 | 0.20 | 0.001 |
| Sex | -0.07 | 0.21 | 0.74 |
| Age | 0.004 | 0.009 | 0.65 |
| BMI | -0.008 | 0.02 | 0.64 |
| **IL-8** |  |  |  |
|  | *B Coefficient* | *Standard Error* | *p-value* |
| SSD diagnosis | 0.07 | 0.26 | 0.78 |
| Sex | -0.07 | 0.26 | 0.80 |
| Age | 0.026 | 0.011 | 0.03 |
| BMI | -0.002 | 0.02 | 0.91 |
| **IL-6** |  |  |  |
|  | *B Coefficient* | *Standard Error* | *p-value* |
| SSD diagnosis | 0.04 | 0.12 | 0.97 |
| Sex | 0.05 | 0.12 | 0.67 |
| Age | 0.009 | 0.005 | 0.10 |
| BMI | 0.009 | 0.01 | 0.37 |
| **IL-4** |  |  |  |
|  | *B Coefficient* | *Standard Error* | *p-value* |
| SSD diagnosis | 0.67 | 0.31 | 0.04 |
| Sex | -0.18 | 0.32 | 0.57 |
| Age | 0.02 | 0.014 | 0.04 |
| BMI | 0.006 | 0.02 | 0.82 |

### 3.3. Plasma Analysis

#### 3.3.1. Cytokine Levels in Plasma by Subject Groups

There were no statistically significant differences in any of the cytokines between SSD and HV in both bivariate and regression analysis after adjusting for sex, age, and BMI (Suppl. Table 1).

### 3.4. Correlations between CSF and blood for all cytokines

No statistically significant correlations were detected in individual cytokines across CSF and plasma when collapsing SSD and HV into one group or when examined groups separately (Suppl. Tables 2 and 3).

## 4. Discussion

In this paper, we report elevated levels of TNFα in CSF in schizophrenia patients compared to healthy volunteers. Importantly, we did not find any significant differences between groups in plasma cytokine levels and did not find any significant correlations between CSF and plasma levels. In addition, we found that female HV had higher CSF TNFα values than male HV but male patients with SZ had higher values than female patients with SZ. Moreover, we found that TNFα and IL-4 were negative correlated with speed of processing and IL-4 was also negative correlated with the overall MCCB score but positive correlated with conceptual disorganization and alogia. Together, these findings provide important insight about cytokine dysregulation in schizophrenia and their potential impact in cognition and language and suggest that peripheral cytokine levels are not reflective of CSF levels.

Our CSF findings contrast with those by Zhu and colleagues (2016) who found decreased CSF TNFα levels in schizophrenia, though their control group consisted of patients with non-suppurative appendicitis instead of healthy volunteers. Although non-suppurative appendicitis reflects a macroscopic local infection, it is possible that these patients manifest a systemic inflammatory response. In addition, the inflammatory response observed in schizophrenia patients does not resemble that associated with an acute infectious disease but rather a chronic low grade inflammatory activation. Although we were able to detect CSF TNFα levels in most of our participants, our results also contrast with Barak et al. (1996) who did not detect CSF TNFα levels using Enzyme link immunoassay (EIA) in neither SZ patients nor in HV. Our findings are in agreement with those by Maxeiner and colleagues (2014) which observed marginally elevated CSF TNFα levels in 16 schizophrenia patients compared to affective disorder patients. Therefore, our data provides support for elevated TNFα as a CSF biomarker that is relevant to the pathophysiology of schizophrenia and potentially serve as diagnostic biomarker or as a predictor of response to treatment in future studies.

The exact mechanism by which TNFα is associated with schizophrenia is unknown. However, it is known that TNFα has two receptor subtypes TNF-R1 and TNF-R2 with the former contributing to neuronal death through the presence of a death domain and the latter associated with neuroprotection (Iosif et al., 2006). In addition, TNFα is known to be secreted by glial cells and regulate synaptic plasticity by inducing glutamatergic activity via an increase in surface α-amino-3-hydroxy5-methyl-4-isoxazolepropionic acid (AMPA) receptors (Beattie et al., 2002). Therefore, elevated CSF TNFα levels may be mechanistically relevant to the dysregulation in glutamate homeostasis associated with schizophrenia (Merritt et al., 2016) and warrants additional investigation.

Our finding that CSF IL-4 was higher in SSD individuals cannot be compared to other studies since no other authors have reported CSF IL-4 levels in schizophrenia. On the other hand, this finding is preliminary since 20 out of 53 data points were imputed. Therefore, additional studies are needed to clarify whether CSF IL-4 levels are abnormal in patients with schizophrenia.

Interestingly, we did not find a significant difference in CSF IL-6 levels between SSD and HV, as has been reported previously by our group and others (Van Kammen et al., 1999, Garver et al., 2003, Soderlund et al., 2009, Sasayama et al., 2013, Hayes et al., 2019, Schwieler et al., 2015 and Coughlin et al., 2016, Gallego et al., 2018), though Barak and colleagues (1995) did not find differences between SZ and HV. Although the reason for these divergent findings is not apparent, all patients in our study were medicated and psychiatrically stable and higher IL-6 levels are observed in acutely relapsed or first episode patients and decrease following antipsychotic treatment (Miller et al., 2011). In our study, however, antipsychotic dose expressed as chlorpromazine equivalents was not correlated with IL-6.

Our finding that female HV have numerically higher CSF TNFα levels compared to HV males is generally consistent with findings that females have a higher innate and adaptive immune response compared to males (Klein et al., 2016). The latter may be due to the higher concentrations of androgens in post-pubertal men which are associated with suppression of immune cell activity (Klein et al, 2016). Interestingly, male patients showed numerically higher TNFα levels compared to female patients, despite HV males having lower TNFα levels than HV females. Unfortunately, no other studies have investigated the impact of sex in CSF TNFα levels. The only two studies that specifically reported sex differences in CSF cytokine levels in SZ found either no differences in IL-6 by sex (Garver et al., 2003) or higher CSF IL-6 levels in both males and female patients compared to male and female controls, with overall higher levels in females compared to males in all strata (Schwieler et al., 2015). Nevertheless, the present findings highlight the importance of investigating the impact of sex on CSF cytokine levels in future studies.

An intriguing finding in our study was that TNFα and IL-4 were negatively correlated with speed of processing (for both NTF and IL-4) and MCCB composite score (for IL-4). In support of this finding, a recent meta-analysis by Patlola and colleagues (2023) reported a significant relationship between inflammatory markers and cognitive dysfunction in schizophrenia, particularly for TNFα and other proinflammatory cytokines. A similar finding was reported by Chukaew and colleagues (2022) who found that serum TNF was negatively correlated to processing speed and attention in a group of chronic schizophrenia patients. However, these findings should be considered preliminary and need to be interpreted with caution given our inability to adjust for multiple comparison due to the sample size of our study, but are useful to generate hypothesis to be tested in future studies.

Strengths of our study include the concurrent collection of blood and CSF samples in a well characterized group of schizophrenia patients and healthy volunteers. Furthermore, our CSF sample collection and processing was standardized since we collected them as part of a research study instead of obtaining them from samples of convenience drawn from clinical practice. Importantly, our findings are not a result of substance use since most subjects had negative urine toxicology tests. On the other hand, our study was limited due to the relatively small sample size, as usually observed in single-site CSF studies, though our sample size was comparable to most prior CSF studies (Gallego et al., 2018). In addition, we lacked smoking data, a variable often associated with increased inflammatory markers. However, we were able to investigate other variables associated with inflammatory markers such as sex, age and BMI, which were appropriately included in the regression model. Finally, we were unable to directly determine the impact of antipsychotic exposure on the outcomes since all patients included in our study were receiving antipsychotic treatment, but we did not find a correlation between TNFα levels and antipsychotic dose expressed as chlorpromazine equivalents.

In summary, the present findings provide important insight concerning cytokine dysregulation in CSF in schizophrenia, particularly of TNFα, and the lack of correlation between CSF and blood cytokine levels. Our data provides initial support for the potential use of CSF TNFα as a biomarker that is potentially relevant to the neuropathophysiology of schizophrenia. Our data also provides additional information about the relationship between cytokine levels and cognitive deficits and provides evidence of the role of sex in immune system dysregulation. Future studies are needed to validate these findings in an independent sample and to determine the impact of antipsychotic exposure e.g., in antipsychotic-naïve or antipsychotic-free participants. In addition, given the challenges in recruiting a large number of individuals for single site CSF studies, subsequent CSF efforts will require multisite studies to increase the probability of finding druggable and reliable biomarkers. Moreover, prospective longitudinal studies that include early psychosis patients are needed to clarify the relationship between CSF TNFα levels and illness progression. Finally, the simultaneous collection of genomic, neuroimaging, and other relevant measures would facilitate the integration and understanding of the inflammatory abnormalities in psychosis across several domains.

## Data Availability

All data produced in the present study are available upon reasonable request to the authors

## Funding

This study was supported in part by a NARSAD Young Investigator Grant (PI: J. Gallego) from the Brain & Behavior Research Foundation, a K23MH100264 from the National Institute of Mental Health (PI: J. Gallego), and R01DK097599 (PI: McNamara). The content is solely the responsibility of the authors and does not necessarily represent the official views of the National Institutes of Health or the Brain & Behavior Research Foundation.

## Declarations of Interest

Drs. Gallego, Castaneda, Jimenez, Alvarez-Lesmes and Lencz have nothing to disclose. Emily Blanco has nothing to disclose. Dr. Malhotra is a consultant to Genomind Inc and InformedDNA,

## Acknowledgements

None

## Notes

### Author Declarations

All subjects provided written informed consent to a protocol approved by the Institutional Review Board of the Northwell Health System

