## Supplementary File for "Evidence for Cytokine Dysregulation in Schizophrenia Spectrum Disorders: A Comparison of Cerebrospinal Fluid and Blood Samples"

**Suppl. Table 1. Plasma Cytokine Levels – Multivariate Linear Regression Analysis**

| **IL-8** | | | |
| --- | --- | --- | --- |
|  | *B Coefficient* | *Standard Error* | *p-value* |
| SSD diagnosis | 1.06 | 0.89 | 0.25 |
| Sex | -1.28 | 0.86 | 0.15 |
| Age | 0.005 | 0.039 | 0.88 |
| BMI | -0.064 | 2.22 | 0.03 |
| **IL-6** | | | |
|  | *B Coefficient* | *Standard Error* | *p-value* |
| SSD diagnosis | -0.03 | 0.45 | 0.95 |
| Sex | -0.67 | 0. | 0.15 |
| Age | 0.005 | 0.039 | 0.88 |
| BMI | -0.064 | 2.22 | 0.03 |
| **IL-4** | | | |
|  | *B Coefficient* | *Standard Error* | *p-value* |
| SSD diagnosis | -0.12 | 1.12 | 0.92 |
| Sex | -1.39 | 1.04 | 0.20 |
| Age | 0.006 | 0.048 | 0.89 |
| BMI | -0.054 | 0.10 | 0.59 |
| **IL-2** | | | |
|  | *B Coefficient* | *Standard Error* | *p-value* |
| SSD diagnosis | 1.05 | 1.15 | 0.36 |
| Sex | -1.69 | 1.10 | 0.14 |
| Age | 0.079 | 0.049 | 0.14 |
| BMI | -0.011 | 0.10 | 0.78 |
| **IL1-ß** | | | |
|  | *B Coefficient* | *Standard Error* | *p-value* |
| SSD diagnosis | 0.093 | 0.46 | 0.84 |
| Sex | -0.76 | 0.45 | 0.1 |
| Age | 0.023 | 0.02 | 0.26 |
| BMI | -0.016 | 0.041 | 0.71 |
| **TNFα** | | | |
|  | *B Coefficient* | *Standard Error* | *p-value* |
| SSD diagnosis | -0.045 | 0.33 | 0.89 |
| Sex | -0.57 | 0.32 | 0.08 |
| Age | 0.0005 | 0.01 | 0.97 |
| BMI | 0.001 | 0.029 | 0.96 |

SSD: Schizophrenia spectrum disorder; BMI: Body mass index

**Suppl. Table 2. Cytokine correlations between CSF and plasma**

| **All subjects (N=39)** | | |
| --- | --- | --- |
| **Cytokine** | **r** | **p-value** |
| TNFα | 0.07 | 0.67 |
| IL-4 | -0.08 | 0.63 |
| IL-6 | 0.19 | 0.23 |
| IL-8 | 0.16 | 0.33 |

r: Pearson correlation coefficient

**Suppl. Table 3. Cytokine correlations between CSF and plasma by subject group**

| **SSD (n=22 )** | | |
| --- | --- | --- |
| **Cytokine** | **r** | **p-value** |
| TNFα | 0.10 | 0.65 |
| IL-4 | -0.001 | 0.99 |
| IL-6 | 0.26 | 0.25 |
| IL-8 | 0.20 | 0.36 |
| **HV (n=17 )** | | |
| **Cytokine** | **r** | **p-value** |
| TNFα | 0.14 | 0.58 |
| IL-4 | -0.18 | 0.49 |
| IL-6 | 0.13 | 0.61 |
| IL-8 | 0.15 | 0.56 |

r: Pearson correlation coefficient

SSD: schizophrenia spectrum disorder

HV: healthy volunteers; r: correlation coefficient
